# Community health workers’ experiences with a digital clinical decision support system for longitudinal hypertension and diabetes care in rural Lesotho: a mixed-methods study

**DOI:** 10.64898/2026.11.03.26367148

**Authors:** Felix Gerber, Fabian Raeber, Motlalentoa Moruthoane, Antony Khaemba, Nitin Bhandari, Lal Kunwar, Giuliana Sanchez-Samaniego, Ravi Gupta, Andréa Williams, Thesar Tahirsylaj, Malebona Mathulise, Thuso Kabi, Mosoetsi Mokaeane, Malehloa Maphenchane, Manthabiseng Molulela, Makhebe Khomolishoele, Mamoronts’ane Pauline Sematle, Lebohang Sao, Mosa Tlahali, Unnati Shankar, Dave Brian Basler, Kevin Kindler, Pauline Grimm, Niklaus Daniel Labhardt, Alain Amstutz

## Abstract

Community health workers (CHWs) are increasingly delivering chronic care in resource-constrained settings but require support to manage complex longitudinal clinical workflows. In the Community-Based chronic Care Lesotho (ComBaCaL) project in rural Lesotho, CHWs provide longitudinal hypertension and diabetes care, supported by the ComBaCaL Community Health Toolkit (CHT) application. The application combines longitudinal participant management and task coordination with clinical decision support for screening, diagnosis, treatment initiation and adjustment, cardiovascular risk management, referral, and follow-up for hypertension and diabetes care. This model of care has demonstrated improved or maintained clinical outcomes compared to facility-based care only. We conducted a mixed-methods study among all 103 participating CHWs to evaluate their experiences with the application.

All CHWs completed a 38-item questionnaire, and 22 participated in qualitative interviews or focus group discussions. Overall usability was high, with a mean System Usability Scale score of 75.6 (SD 13.0), and 98 (95%) CHWs were satisfied or very satisfied with the application. General application functions and diagnosis and treatment tasks were rated easy or very easy by 98% of participants. The most frequently reported benefits were task reminders (92%), structured clinical guidance (87%), overview of allocated participants (81%), and retention of participant information (81%). Main challenges were lengthy questionnaires, difficulties charging tablets, and limited internet connectivity. Qualitative findings showed that ease of use increased with experience, supported by training and supervision. Participants described how algorithmic guidance and task reminders supported care delivery, while also identifying tensions between standardized workflows and flexibility in day-to-day care.

The ComBaCaL CHT application was highly usable and valued by CHWs. High perceived usability extended from general application functions to complex clinical workflows. CHT demonstrated suitability as a configurable framework for implementing such context-specific workflows for hypertension and diabetes care when accompanied by appropriate workflow design, training, and supervision.

**Author summary:** High blood pressure and diabetes require repeated follow-up and treatment decisions, which is difficult where healthcare professionals are scarce. Community health workers (CHWs) may help expand access to this care, but they need appropriate tools to manage complex clinical tasks. We evaluated a digital application designed to support CHWs in delivering hypertension and diabetes care in rural Lesotho. Overall, 103 CHWs used an application built with the Community Health Toolkit to support hypertension and diabetes care. The application helped manage participants and follow-up tasks and provided step-by-step guidance for screening, diagnosis, treatment, referral, and repeated care visits. All 103 CHWs completed a survey, and 22 participated in interviews or focus group discussions.

CHWs found the application easy to use and useful for their work. Frequently reported benefits included reminders, procedural guidance, better participant oversight, and retention of participant information over time. Ease of use improved with experience and was supported by training and supervision. Challenges included lengthy research questionnaires, difficulties charging tablets, limited internet connectivity, and standardized digital workflows that did not fit day-to-day circumstances.

Our findings show that a configurable open-source digital application can support CHWs in delivering complex, longitudinal hypertension and diabetes care in a remote rural setting.

## Introduction

Task-shifting to lay health providers such as community health workers (CHWs) has emerged as a promising strategy to improve access to essential health services in resource-constrained settings [1,2]. CHWs are trained members of their communities and can help overcome barriers related to travel distance, costs, and healthcare workforce shortages. While CHW-led healthcare delivery models have demonstrated effectiveness for health promotion, screening, and linkage to care, their role in longitudinal management of chronic diseases, such as hypertension and type 2 diabetes, remains limited [3,4].

Hypertension and type 2 diabetes are among the leading contributors to morbidity and mortality in low-and middle-income countries, yet gaps in diagnosis, treatment, and long-term control persist, particularly in rural areas [5–7]. Shortages of trained healthcare professionals, geographic barriers to care, and limited health system resources continue to constrain access to care for these conditions [8]. Their management requires repeated assessments, treatment decisions, medication management, and long-term follow-up, activities that have traditionally been performed by more highly trained healthcare professionals. Expanding the role of CHWs to include these tasks therefore requires approaches that ensure care remains safe, consistent, and aligned with clinical guidelines.

Digital clinical decision support systems (CDSSs) have been proposed to extend the scope and quality of community-based healthcare delivery [9]. By guiding users through standardized workflows, supporting clinical decision-making, and enabling remote supervision, digital tools may help address some of the operational and clinical challenges associated with task-shifted care [10]. Additionally, many community health programs continue to rely on fragmented or paper-based information systems, making it difficult to maintain longitudinal patient records, coordinate follow-up, and provide effective supervision [11]. Digital platforms that combine longitudinal data capture with clinical decision support could thus support both continuity of care and program oversight. However, evidence on how CHWs experience and use digital systems that support complex care processes, including diagnosis, treatment initiation, titration and long-term follow-up remains limited.

Many digital health interventions are developed as stand-alone applications with limited interoperability, scalability, and long-term sustainability [12]. Increasing attention is therefore being directed toward digital public goods and open-source platforms that can be adapted to local contexts while supporting integration into routine health systems [13]. The Community Health Toolkit (CHT) is an established open-source framework for developing configurable digital health applications for community health systems. Its core framework provides a common user interface that can be customized to support different health system hierarchies, roles, and program needs. Designed as an offline-first tool, CHT supports longitudinal patient and household records, care guides for health-worker decision support, tasks to ensure timely care, in-app dashboards for performance management, and messaging for rapid communication [14]. CHT-based applications have been implemented across diverse health programs and on a national scale in several countries. Previous studies of CHT-based applications have demonstrated usability and acceptability for more focused workflows, including two-way communication and follow-up after voluntary medical male circumcision and for retention in HIV care [15,16]. Evidence remains limited, however, on the use of CHT to support the complex, longitudinal management of hypertension and type 2 diabetes by lay CHWs.

The Community-Based chronic Care Lesotho (ComBaCaL) project was established in rural Lesotho to develop and evaluate innovative models of community-based chronic care [17]. Within the project, a mobile application based on the CHT framework was developed to support longitudinal population-based data collection alongside screening, diagnosis, treatment initiation, medication management, and longitudinal follow-up of individuals with hypertension and type 2 diabetes (ComBaCaL CHT application) [18–20]. The feasibility and acceptability of the approach were first assessed in a pilot study conducted in ten villages with one CHW each [21–23]. Lessons learned during this pilot informed further refinement of the application and its workflows, most notably the consolidation of separate screening, treatment-initiation, and follow-up forms into a single, adaptive form that adjusts to each participant’s current diagnostic and treatment status, accommodating any care state encountered over repeated visits. The resulting system was subsequently implemented by 103 CHWs across 103 rural villages and formed a core component of three cluster-randomized trials evaluating CHW-led hypertension and diabetes care [18,19]. These trials demonstrated improved hypertension control among individuals with uncontrolled hypertension, improved engagement in care among individuals with type 2 diabetes, and non-inferior outcomes among individuals with controlled hypertension receiving the additional option of CHW-led compared with facility-based nurse-led care alone [24–26]. While the previous studies examined the initial feasibility and design of the digital approach and subsequent trials established the effectiveness of the broader care models, the usability of the mature application and CHWs’ experiences using it for longitudinal care have not been comprehensively evaluated.

Understanding how frontline health workers experience digital CDSSs is essential for informing future scale-up and adaptation of community-based care models [27]. We therefore describe the design and implementation of the ComBaCaL CHT application and report a mixed-methods evaluation among all 103 CHWs using the system. We aimed to assess its usability, user satisfaction, and perceived value and to explore how CHWs experienced the use of the platform to support complex longitudinal hypertension and diabetes care in remote rural communities.

## Materials and methods

### Study setting

The ComBaCaL project is a community-based research and service delivery platform established by a consortium including the Lesotho Ministry of Health, local community representatives, the local non-profit organization SolidarMed and the University of Basel, Switzerland, in the rural districts of Butha-Buthe and Mokhotlong, Lesotho. Its core component is a population-based cohort comprising 103 villages with one participating CHW per village. Villages were selected independently of CHW characteristics; where no active CHW was available, a new CHW was elected by the village population in collaboration with the Ministry of Health according to national procedures [28]. The cohort serves as a platform for the evaluation of community-based care models through embedded cluster-randomized trials and implementation studies. At the time of this evaluation, the primary analysis of three cluster-randomized trials assessing CHW-led hypertension and diabetes care had been completed [24–26]. All CHWs were provided with Android tablets (Lenovo TabM10 FHD Plus Model TB-X606X) with the ComBaCaL CHT application installed and with power banks; CHWs in selected villages with limited access to electricity additionally received portable solar charging systems.

After cohort enrollment and baseline data collection, all CHWs conducted community-based screening and diagnosis of hypertension and type 2 diabetes. In 51 of 103 villages allocated to the intervention arms of the nested trials, CHWs additionally deliver protocol-based treatment and follow-up of eligible participants with hypertension and/or type 2 diabetes. CHWs received structured initial training on the clinical protocols and the application over roughly 10 days, followed by short refresher trainings at roughly 6-month intervals. During implementation, they received clinical support from routine facility-based nursing staff during regular monthly meetings and by study staff who reviewed submitted records and program-monitoring dashboards through the web-based interface and provided clinical and operational feedback. Technical support for devices, connectivity, and application issues was provided by the ComBaCaL study team through phone calls or village visits if needed. Details of the ComBaCaL cohort, trial designs, intervention protocols and consent procedures have been published previously [17–20,24–26].

### Application design and functionality

To support longitudinal population-based data collection and community-based hypertension and diabetes care, we developed the ComBaCaL application (hereafter referred to as “the application”) using the open-source CHT Core Framework (versions 3.12, 3.14, 4.6, and 4.13), available at https://github.com/clinepi-usb/cht-combacal. The CHT Core Framework provided the common user interface and underlying architecture for user roles and hierarchies, household and participant records, longitudinal participant profiles, task management, reporting, program monitoring, participant search, and offline data storage and synchronization. Within this framework, we configured the ComBaCaL-specific forms, task rules, research and cohort workflows, translations, and clinical decision support for hypertension and diabetes care.

The CDSS component provided protocol-based guidance for screening, diagnosis, treatment initiation and adjustment, cardiovascular risk management, referral, counseling, and longitudinal follow-up. A messaging component available within the CHT Core Framework was not implemented because it was not a key project requirement and integration with local short message service (SMS) infrastructure proved technically challenging. The platform further supports role-based access through configurable user hierarchies. CHWs access the application on the password-protected tablets, while supervisors access the same system through a web-based interface that allows review of participant records, submitted reports, and program monitoring information across all villages under their supervision. Data were stored locally on the tablets for offline use and synchronized when connectivity was available to secure servers hosted at the University Hospital Basel, Switzerland. Routine health-facility staff continued to use existing facility-based documentation and reporting systems. Although CHT supports interoperability with other complementary health-information systems, direct electronic integration with the national reporting architecture was outside the scope of this initial research implementation. To maintain alignment with established Ministry of Health reporting pathways, CHWs reported relevant activities to supervising facility staff for entry into routine facility registers rather than replacing the existing reporting architecture with direct digital reporting from the application.

The application workflow begins with household registration followed by registration of individual household members. After informed consent and baseline data collection, participants become eligible for hypertension and type 2 diabetes screening according to predefined criteria. Screening and diagnosis followed the Lesotho Standard Treatment Guidelines with confirmation of elevated blood pressure or blood glucose required on two different visits for a diagnosis [29]. Participants remained within a longitudinal record that was updated during subsequent encounters, with previous and current clinical information used to determine subsequent assessments and care. Full screening and diagnostic algorithms are provided in the Supporting Information (S1 and S2 Figs).

During screening and follow-up, CHWs collected information on medication use, comorbidities, symptoms, blood pressure, blood glucose, and other clinical information required to determine the participant’s current care status. In intervention villages, the CDSS component additionally guided protocol-based treatment and follow-up. This included lifestyle counseling, initiation, titration and de-escalation of first-line pharmacological treatment (amlodipine/hydrochlorothiazide combination pill for hypertension; metformin for type 2 diabetes), cardiovascular risk-based preventive treatment (atorvastatin and aspirin according to predefined eligibility criteria), referral, and follow-up scheduling. In control villages, CHWs provided lifestyle counseling and referral to facility-based care. Full treatment algorithms are provided in the Supporting Information (S3 and S4 Figs).

Unlike its pilot version, which used separate forms for screening, diagnosis, treatment initiation, and follow-up, the final application is designed as a longitudinal, adaptive workflow [21,22]. This redesign was necessary because participants frequently transition between different care states over time, making linear screening-to-treatment workflows insufficient for routine chronic disease care. The application integrates information from previous visits and current assessments to determine a participant’s care status and dynamically guide the CHW through the next appropriate action (Fig 1). This design allows a single workflow to support the full continuum of community-based chronic disease care while accommodating changes in clinical circumstances, participant preferences, treatment uptake, equipment availability, medication availability, and other factors encountered during routine care.

**Fig 1.**
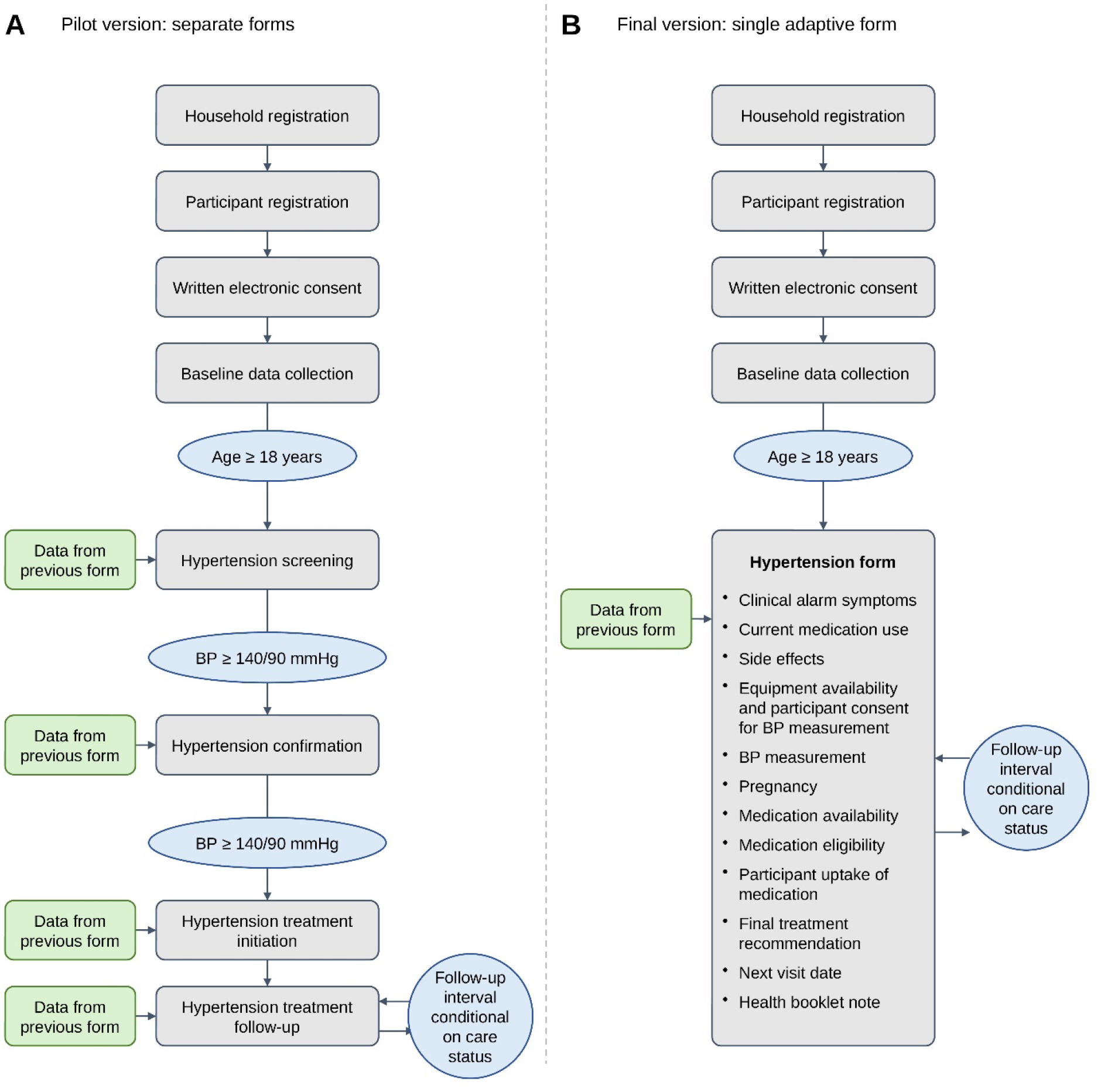
Simplified overview of the ComBaCaL application workflow. Each grey box represents a form within the application. Following registration and baseline assessment, disease-specific workflows are triggered according to predefined eligibility criteria. Information from previous encounters and data entered during the current visit are integrated to determine the participant’s current care status, generate management recommendations, and schedule subsequent follow-up activities. Panel A: Workflow of the pilot version with different forms for screening, confirmation of diagnosis, treatment initiation and follow-up. Panel B: Workflow of the final version with a single adaptive form that integrates relevant information from previous and current encounters.

### Study design

We conducted a mixed-methods study to evaluate CHWs’ experiences using the application. The study combined a quantitative survey administered to all participating CHWs with qualitative individual in-depth interviews (IDIs) and focus group discussions (FGDs) among a purposively selected subset of CHWs. Quantitative and qualitative data were analyzed separately and subsequently integrated during interpretation.

### Quantitative data collection

All 103 CHWs participating in the ComBaCaL project were invited to complete a structured 38-item survey. The survey collected information on demographic characteristics, education level, English proficiency, and prior digital experience. Participants were additionally asked about usability, satisfaction, perceived value, ease of performing specific clinical tasks, perceived benefits and challenges of system use, and recommendations for future implementation.

Usability was assessed using the System Usability Scale (SUS), a validated ten-item questionnaire consisting of alternating positive and negative statements rated on a five-point Likert scale ranging from strongly disagree to strongly agree [30]. Responses were converted into a composite score ranging from 0 to 100 according to standard scoring procedures, with higher scores indicating greater perceived usability.

Additional survey items were adapted from established instruments and tailored to the ComBaCaL context. Items assessing perceived usefulness, communication, interface, and overall satisfaction were adapted from the mHealth App Usability Questionnaire (MAUQ) [31], while selected items on perceived effects on care delivery and clinical decision-making were adapted from the Primary Care Information Project Provider Survey [32]. Task-specific ease of use was assessed using questions adapted from the Single Ease Question approach [33], rated on a five-point scale from very easy to very hard. Additional study-specific questions assessed perceived benefits and challenges of application use and suggestions for future implementation. Survey questions and answer choices were presented in both English and the local language, Sesotho. The complete survey instrument is provided in the Supporting Information (S1 Appendix).

### Qualitative data collection

To obtain an in-depth understanding of CHWs’ experiences using the application, we conducted semi-structured IDIs and FGDs. Participants were selected using purposive maximum-variation sampling to capture a broad range of experiences across districts, age, sex, village accessibility, trial arm allocation, digital literacy, and previous work experience as a CHW. The qualitative sample size was determined a priori through purposive maximum-variation sampling, with the option to conduct additional interviews had the predefined sample not proven sufficiently rich; the sampled range was judged to provide comprehensive accounts, and no further interviews were required. Selected CHWs were invited face-to-face during a regular clinical training, and all invited participants agreed to participate. Four IDIs and two FGDs involving a total of 22 CHWs were conducted. IDIs included two participants from Butha-Buthe district and two participants from Mokhotlong district. The FGDs included eight participants in Butha-Buthe and ten participants in Mokhotlong. Four participants had previous experience working as CHWs before the introduction of the digital system and were therefore able to reflect on differences between paper-based and digital workflows.

Semi-structured interview guides explored experiences using the application, perceived benefits and challenges, training and support needs, usability, and the role of the application in supporting delivery of hypertension and diabetes care. The guides were pilot-tested before data collection. IDIs and FGDs were conducted in Sesotho by Motlalentoa Moruthoane, BSc Social Sciences, a male trained qualitative research assistant with previous experience conducting qualitative interviews and FGDs. He was not involved in delivering the intervention or supervising participating CHWs. Participants were informed of his role and the purpose of the qualitative study before data collection. IDIs and FGDs were conducted at the training venue, with only the researcher and participants present, and lasted approximately 30–40 and 90–100 minutes, respectively. Sessions were audio-recorded and translated into English during transcription. No repeat interviews were conducted and no field notes were taken. Transcripts were not returned to participants for comment or correction, and formal participant checking of the findings was not undertaken. The semi-structured guide for IDIs and FGDs is available in Supporting Information (S2 Appendix).

### Quantitative analysis

Survey responses were analyzed descriptively using R version 4.3.3 [34]. Continuous variables were summarized using means and standard deviations or medians and interquartile ranges, as appropriate. Categorical variables were summarized using frequencies and percentages. SUS scores were calculated according to standard scoring procedures [35]. Responses to individual SUS items, perceived value and satisfaction measures, ease-of-task assessments, perceived benefits, perceived challenges, and recommendation questions were summarized descriptively.

### Qualitative analysis

Qualitative data were analyzed using framework analysis. IDI and FGD transcripts were read repeatedly to achieve familiarity with the data. An initial analytical framework was informed by domains covered in the quantitative survey and semi-structured interview guide, including usability and perceived value, ease of use of general application functions and clinical tasks, and perceived benefits, challenges, and opportunities for improvement. Coding combined deductive application of these predefined domains with inductive identification of additional codes arising from the data. Two researchers (AA, FG) conducted the analysis collaboratively and iteratively. An initial coding framework and analysis matrix were developed, compared across transcripts, discussed, and refined until agreement was reached. Codes and categories were further refined through comparison across participants and data sources and organized into four overarching themes: (1) usability, learning, and enabling conditions; (2) how the application supported longitudinal chronic care; (3) challenges and tensions; and (4) suggested adaptations and future development. Findings extending beyond the predefined survey domains were retained within the analytical framework. Coding and matrix development were performed manually. Representative quotations were selected to illustrate the findings. Quotations are presented from transcripts translated from Sesotho into English, with minor edits for readability. The analysis matrix is outlined in S1 Table. The study is reported in accordance with the Consolidated Criteria for Reporting Qualitative Research (COREQ) [36]; the completed checklist is presented in the Supporting Information (S1 Checklist).

### Integration of quantitative and qualitative findings

We used a convergent mixed-methods design in which quantitative and qualitative data were analyzed separately and integrated during interpretation and reporting. Because the qualitative interview guides covered domains also assessed in the survey, the two data sources addressed overlapping aspects of usability, clinical task support, perceived benefits, and implementation challenges. During integration, qualitative findings were compared with the corresponding survey results to identify areas of convergence, divergence, and additional insights not captured by the predefined survey items. Qualitative findings were used both to contextualize quantitative results and to explore mechanisms through which the application supported hypertension and diabetes care. Reporting of the mixed-methods design follows the Good Reporting of A Mixed Methods Study (GRAMMS) criteria [37]; the completed checklist is provided in the Supporting Information (S2 Checklist).

### Ethics

The ComBaCaL cohort study and ongoing nested trials, including the present usability sub-study, were approved by the National Health Research Ethics Committee in Lesotho (ID 210-2022 and ID 210-2022 Nested). The Ethikkommission Nordwestschweiz (EKNZ) in Switzerland issued declarations that the study met all requirements for a Swiss research study (AO_2022-00058, AO_2022-00074 and AO_2022-00077). The nested trials are registered on ClinicalTrials.gov (NCT05596773, NCT05743387, and NCT05684055). The study was conducted in accordance with the principles of the Declaration of Helsinki. All participants provided written informed consent before participation. Additional information regarding the ethical, cultural, and scientific considerations specific to inclusivity in global research is included in the Supporting Information (S3 Checklist).

### Patient and public involvement

The Ministry of Health of Lesotho and the District Management Teams of Butha-Buthe district were among the initial members of the research consortium. Community-engaged research is key in the ComBaCaL project. The CHWs are members of the community and have been elected by the community. Research questions for nested trials are based on feedback from community-members during implementation of the ComBaCaL activities. Prior to implementation of the ComBaCaL cohort, we conducted a pilot study to incorporate views and perceptions of the local community and CHWs in the final design of the study and nested trials. Community representatives will be involved in the development of protocols for further nested trials.

## Results

### Participant characteristics and implementation exposure

All 103 CHWs participating in the project completed the survey between November 24 and December 17, 2024. During the same period, 22 CHWs participated in the qualitative component through four IDIs and two FGDs. All 103 CHWs had used the application for participant registration, screening, and diagnosis of hypertension and type 2 diabetes, while the 51 CHWs in intervention villages had additionally used it to support protocol-based treatment and longitudinal follow-up. By the time of the evaluation, the application had been used to register 5283 households and 16297 participants, screen 8096 adults for hypertension and 6135 adults for type 2 diabetes, and support longitudinal care for 401 participants with hypertension and 60 participants with type 2 diabetes enrolled in the nested cluster-randomized trials. The median age of participating CHWs was 32 years (IQR 28–39), 74 (72%) were female, and 94 (91%) had completed at least secondary education. Although 88 (85%) reported previous experience using a smartphone or tablet, 15 (15%) had no prior experience with either device (Table 1).

**Table 1.** Characteristics of participating community health workers.

| **Characteristic** | **Community health workers (N=103)** |
|---|---|
| **Age, years, median (IQR)** | **32 (28–39)** |
| **Sex, n (%)** |  |
| Female | 74 (72%) |
| Male | 29 (28%) |
| **District, n (%)** |  |
| Butha-Buthe | 52 (50%) |
| Mokhotlong | 51 (50%) |
| **Trial arm, n (%)** |  |
| Control | 52 (50%) |
| Intervention | 51 (50%) |
| **Education, n (%)** |  |
| Primary school | 9 (9%) |
| Secondary school | 74 (72%) |
| Tertiary education | 20 (19%) |
| **Self-rated English proficiency, n (%)** |  |
| Poor | 2 (2%) |
| Moderate | 45 (44%) |
| Good | 31 (30%) |
| Very good | 25 (24%) |
| **Prior digital experience, n (%)** |  |
| No previous smartphone/tablet use | 15 (15%) |
| Previous smartphone/tablet use | 88 (85%) |

IQR, interquartile range.

The 22 CHWs in the qualitative component comprised four IDI participants (two per district, aged 24–36) and 18 FGD participants across two groups (Butha-Buthe: eight CHWs, aged 25–39; Mokhotlong: ten CHWs, aged 23–44).

### Overall usability, perceived value and satisfaction

Overall usability, perceived value and satisfaction with the application were high (Table 2; n=103). The mean SUS score was 75.6 (SD 13.0). Almost all CHWs rated the application as useful for their work (102, 99%) and reported that it made service provision easier (102, 99%), with similarly high agreement that it improved patient communication, clinical decision-making, and quality of care. Satisfaction and endorsement were comparable: 98 (95%) were satisfied or very satisfied, 100 (97%) considered it useful for all CHWs, and 97 (94%) agreed that its benefits outweighed its challenges.

**Table 2.** Usability, perceived value and satisfaction with the ComBaCaL application.

| **Measure** | **Community health workers (N=103)** |
|---|---|
| SUS score, mean (SD) | 75.6 (13.0) |
| SUS score, median (IQR) | 77.5 (67.5–85.0) |
| Application useful for work, n (%) | 102 (99%) |
| Application makes service provision easier, n (%) | 102 (99%) |
| Application improves patient communication, n (%) | 102 (99%) |
| Application improves clinical decision-making, n (%) | 101 (98%) |
| Application improves quality of care, n (%) | 102 (99%) |
| Benefits of the application outweigh the challenges, n (%) | 97 (94%) |
| Satisfied or very satisfied with application, n (%) | 98 (95%) |
| Application would be useful for all CHWs, n (%) | 100 (97%) |

CHW, community health worker; IQR, interquartile range; SD, standard deviation; SUS, System Usability Scale (score range 0–100; higher scores indicate better usability). Agreement items were rated on a five-point scale from strongly disagree to strongly agree and compared to a paper-based system; values show the number (%) who agreed or strongly agreed.

At the SUS item level (S2 Table), responses were strongly favorable for frequent use, integration, and confidence: 98% would like to use the application frequently, 96% found its functions well integrated, and 96% felt confident using it, while most disagreed that it was complex (83%) or awkward to use (85%). Responses were more mixed on the learnability items: while 74% expected most CHWs to learn the application quickly, a majority reported needing to learn a lot of things before getting started (61%) and 31% reported potentially needing technical support. Only 32% responded favorably to the item on perceived inconsistency, compared with 96% who reported that the application’s functions were well integrated.

The qualitative data contextualized these findings by showing that ease of use developed with experience. Participants described initial difficulty during training and early use, followed by increasing familiarity with the application:

> *“When I first used CHT I thought it was a lot during the training when we were practicing, but as I was using it, it became easy. Although it gets easy with time, you still forget other things, therefore you need to keep reminding yourself all the time like today at the refreshers.”* (IDI, female CHW, Butha-Buthe)

This trajectory extended to CHWs with no prior smartphone experience, consistent with the expectation that most would learn the application quickly:

> *“It was my first time using a smartphone, it was so difficult to navigate. But now it’s easy.”* (FGD, Mokhotlong)

Other participants similarly described practice during training and access to supervisors as helping them become comfortable with the system, while noting that some CHWs continued to require additional training. The bilingual English/Sesotho design was also described as facilitating use and supporting the application’s fit with CHWs’ work. At the same time, participants raised concerns about whether the same experience would extend to the wider CHW workforce, particularly older or less educated CHWs with limited digital experience. One CHW explained:

> *“It could be useful but those CHWs are old people most of them and did not really go far in terms of education. It might be really complicated to train them to use CHT.”* (IDI, male CHW, Butha-Buthe)

This concern was not universal: other participants considered such barriers potentially manageable with adequate training and support.

### Ease of use of general application functions and clinical tasks

CHWs reported that both the core application functions and clinical tasks were easy to perform using the application (Table 3). Registering households and participants, keeping track of pending tasks, and reviewing submitted reports were rated easy or very easy by almost all participants (94–97%), as were diagnosis of hypertension and type 2 diabetes (98%) and giving correct treatment advice (94%).

**Table 3.** Ease of use of general application functions and clinical tasks and perceived added value of the application.

| **Task** | **Community health workers (N=103), n (%)** |
|---|---|
| **General application functions<sup>a</sup>** | **Easy or very easy** |
| Registering new households or participants | 100 (97%) |
| Keeping overview of pending tasks | 100 (97%) |
| Checking and editing submitted reports | 97 (94%) |
| **Clinical tasks<sup>a</sup>** | **Easy or very easy** |
| Diagnosing hypertension and type 2 diabetes | 101 (98%) |
| Providing hypertension and type 2 diabetes treatment<sup>b</sup> | 48 (94%) |
| **Perceived added value of the application<sup>c</sup>** | **Much or somewhat more difficult** |
| Diagnosing hypertension and type 2 diabetes without the application | 66 (64%) |
| Providing hypertension and type 2 diabetes treatment without the application<sup>b</sup> | 38 (75%) |

<sup>a</sup>Assessed on a five-point Likert scale from very easy to very hard.
<sup>b</sup>Only the 51 CHWs in the intervention arms (percentages based on n=51).
<sup>c</sup>Assessed on a five-point Likert scale from much easier to much more difficult.

The application was also seen as adding value to clinical work: 66 (64%) CHWs reported that diagnosing hypertension and diabetes without it would be more difficult, and 38 of 51 (75%) intervention CHWs reported that providing hypertension and type 2 diabetes treatment would be more difficult without the application.

Qualitative findings further illustrated the perceived added value of the application for clinical tasks. Participants described the application as providing step-by-step guidance through screening, diagnosis, and treatment and indicating the next appropriate action:

> *“It is very helpful because after screen I write the results in the CHT and automatically it tells me what to do next. It simplifies my job.”* (IDI, female CHW, Mokhotlong)

Participants also described built-in safeguards, including warnings for potentially incorrect entries or medication doses, contraindication alerts, and checks against medication recorded at previous visits. One CHW explained:

> *“For example, if a client has liver problems it will tell me that this person is not supposed to take these pills. This would put us in jeopardy of putting people on treatment who are not supposed to be.”* (IDI, female CHW, Mokhotlong)

### Perceived benefits, challenges and opportunities for improvement

The most frequently reported benefits of the application were task reminders (92%), guidance through procedures and questionnaires (87%), an improved overview of participants (81%), and secure storage and retention of participant information (81%) (Table 4). Remote supervisor checking was reported to increase confidence by 77 (75%) CHWs, while 75 (73%) reported greater confidence that messages communicated to participants were correct.

**Table 4.** Perceived benefits and challenges and suggested improvements of the clinical decision support system.

| **Measure** | **Community health workers (N=103), n (%)** |
|---|---|
| **Perceived benefits** |  |
| Task reminders | 95 (92%) |
| Guidance for procedures and questionnaires | 90 (87%) |
| Overview of participants and tasks | 83 (81%) |
| Data saved on tablet and record retention | 83 (81%) |
| Remote supervisor checking gives confidence | 77 (75%) |
| Confidence that messages to participants are correct | 75 (73%) |
| Access to participant information | 67 (65%) |
| Other benefit | 13 (13%) |
| **Perceived challenges** |  |
| Questionnaires too long | 66 (64%) |
| Charging the tablet difficult | 50 (49%) |
| Poor internet connection makes syncing difficult | 34 (33%) |
| Takes too much time | 21 (20%) |
| Cannot adapt tasks to own schedule | 16 (16%) |
| Too many technical problems | 14 (14%) |
| Too many tasks | 13 (13%) |
| Supervisors can check entered data | 6 (6%) |
| Data privacy concerns | 5 (5%) |
| Too complicated to use | 2 (2%) |
| Other challenge | 2 (2%) |
| **Suggested improvements** |  |
| Better overview of households and participants | 51 (50%) |
| SMS messages to clients | 48 (47%) |
| Fewer questions | 34 (33%) |
| More detailed information/instruction | 31 (30%) |
| Ability to schedule tasks themselves | 25 (24%) |
| Less detailed information/instruction | 18 (17%) |
| Other improvement | 4 (4%) |

SMS, short message service. Participants could select more than one option in each section.

Qualitative findings illustrated how these benefits supported day-to-day work. Participants described task reminders as helping them plan upcoming visits and prepare required medicines and other commodities, while longitudinal participant records facilitated access to information from previous encounters and reduced reliance on paper records that could be lost. Participants also linked the clinical guidance provided by the application to greater confidence and credibility with patients. Beyond these direct functions, some described digital documentation as strengthening accountability by making completed work traceable:

> *“Without CHT I think our work would lack credibility, there was going to be a lot of cheating but with CHT there is no chance of cheating.”* (FGD, Butha-Buthe)

Another participant similarly contrasted the application with paper-based reporting, where tasks could be recorded as completed without necessarily having been performed. While supervisory visibility was frequently perceived positively, 6 (6%) CHWs identified supervisors being able to check entered data as a challenge and 5 (5%) reported data-privacy concerns (Table 4). Qualitative accounts also showed that task visibility could have unintended consequences when completion depended on participant availability: one participant noted that tasks remaining pending could make it appear that the CHW was not doing the work.

The most frequently reported challenges were long questionnaires (64%), difficulty charging tablets (49%), and limited internet connectivity (33%). Qualitative accounts indicated that much of the questionnaire burden related to research-specific consent and baseline data collection rather than the clinical care workflows themselves. Some participants described these questions as lengthy or intrusive. Charging difficulties were particularly relevant in villages without electricity, while poor network connectivity primarily affected synchronization. Participants also described occasional tensions between following standardized decision support and responding to situations in which the application’s recommendation did not align with their expectations or with care provided elsewhere.

> *“That causes a lot of confusion because as a person you would also want to work the way you were trained. But at the end of the day CHT forces you to do what it says.*” (IDI, female CHW, Butha-Buthe)

The participant clarified that this did not occur often but described situations in which the algorithm’s recommendation differed from subsequent facility treatment. A related tension arose when the algorithm recommended reducing or discontinuing antihypertensive medication. Participants described difficulty explaining this to patients who understood antihypertensive treatment to be lifelong:

> *“…some of the patients do not trust what we say to them because they have an old information that when you are on this kind of treatment you are not to stop taking medication.”* (FGD, Mokhotlong)

The same participant suggested that confirmation from a facility nurse could help patients accept such treatment changes. In one interview, a CHW also described having become reliant on the application’s confirmation when diagnosing hypertension and type 2 diabetes, illustrating that decision support could increase confidence while potentially creating dependence on the system.

Participants also identified challenges related to information continuity across care providers. They described patients using multiple personal health booklets, treatment changes made at health facilities that were not always known to the CHW, and medication obtained from multiple sources, creating a potential risk of duplication.

> *“A challenge of clients who mix pills or overdose because they will be taking medication from me and when the outreach is there, they also go there and get medication*.” (FGD, Butha-Buthe)

Suggested improvements included a better overview of households and participants (50%), automated SMS reminders for participants (47%), fewer questions (33%), and greater flexibility in scheduling tasks (24%). The qualitative findings supported the desire for greater scheduling flexibility: participants valued automated task reminders but described situations in which predefined task timing did not align with participant availability, and some suggested being able to schedule their own visits while retaining supervisory visibility. Although SMS reminders were among the most requested additions, qualitative accounts revealed divergent views. Some participants considered SMS useful for reminding participants of upcoming visits, whereas others raised concerns about limited phone ownership, low literacy, confidentiality on shared phones, and potential family conflict or safety concerns:

> *“Most of our patients don’t have phones, they share phones. Imagine now sending a text message, it is going to raise a lot of suspicions.”* (FGD, Mokhotlong)

The broader discussion in the same FGD included concerns about disclosure of illness to family members and potential conflict arising from messages sent between CHWs and participants. One interview participant suggested telephone calls as a preferable alternative for some participants. Additional suggestions arising from the qualitative data included enabling consultation with supervisors directly within the application, extending digital workflows to health facilities and routine Ministry of Health reporting, and broadening the platform to additional community health services such as HIV testing and family planning.

## Discussion

In this mixed-methods evaluation among all 103 CHWs delivering community-based hypertension and type 2 diabetes care in rural Lesotho, the ComBaCaL application was highly usable and valued by its users. CHWs rated both the general application functions, such as participant registration and overview, task management and reporting, and the more demanding clinical tasks of diagnosis and treatment as easy, while most reported these would be harder to perform without the application. The qualitative findings extended the survey results: usability developed through experience and was sustained by training and supervision, and CHWs described tensions around standardization, operational flexibility, accountability, and continuity of care between community and facility levels. Together with the three nested cluster-randomized trials, which provided clinical evidence for the broader CHW-led care models, this study provides an additional, user-centered account of how the underlying application was experienced [24–26].

Digital CDSSs have enabled several task-shared models of non-communicable disease care, but in almost all of them the authority to initiate and adjust treatment has remained with a professional cadre rather than the CHW [38]. In SMARThealth India, CHWs conducted tablet-based cardiovascular risk assessment and referral while the decision support and treatment decisions were directed to primary-care physicians [39]. In more recent work in Kenya and Uganda, CHWs facilitated home-based hypertension care, but titration was clinician-driven through telehealth [40], and in Guatemala CHWs used a smartphone application to screen for and diagnose hypertension and diabetes, with treatment adjustment made by a healthcare professional [41,42]. While mWellcare, a tablet-based system for integrated hypertension and diabetes management, delivered a comparably broad care support, it was nurse-led rather than delivered by lay workers [43]. The ComBaCaL model extends this experience by enabling CHWs themselves to deliver the full longitudinal pathway at community level — diagnosis, independent treatment initiation and titration, cardiovascular risk management, referral and repeated follow-up.

The good overall usability, reflected in a mean SUS score of 75.6, above the commonly cited benchmark of 68 [30], is notable given the clinical and longitudinal complexity of the application. The common interface, longitudinal records, task management, and offline-first architecture were provided by the established CHT Core Framework, while ComBaCaL-specific workflows and clinical decision support were configured within it. The observed usability therefore reflects the combination of the underlying framework, application configuration, and the training and supervision provided, consistent with evidence that enabling conditions are important for health workers’ adoption of digital technologies in low- and middle-income settings [44]. At the item level, responses were strongly favorable for integration and confidence, while learnability items were more mixed, corresponding with qualitative accounts of initial difficulty followed by increasing confidence with practice. Responses to the single reverse-worded item on inconsistency were a notable exception: only a minority responded favorably, despite near-universal agreement that the application’s functions were well integrated. This most likely reflects variable interpretation due to lack of a specific term in Sesotho rather than genuinely inconsistent performance.

The use of CHT is also relevant to the potential for wider implementation. Rather than developing a stand-alone application specifically for this project, the ComBaCaL workflows were configured within an open-source framework designed for reuse and large-scale deployment and already used in large-scale and national community health programs in multiple countries [14]. This does not establish the scalability or sustainability of the ComBaCaL application itself, but it provides an established technical foundation on which the application can be adapted and extended without rebuilding the underlying architecture.

Beyond overall usability, the findings illustrate how the application supported the longitudinal demands of chronic care. Most CHWs reported that diagnosis and treatment would be more difficult without the application, and qualitative accounts linked this support to step-by-step guidance, safeguards, access to information from previous encounters, and task reminders. The workflow carried information forward between visits and adapted the next recommended action as clinical status, treatment uptake, medication availability, participant preferences, or referral needs changed over time. This allowed screening, diagnosis, treatment, and follow-up to be managed within a single longitudinal pathway. Taken together, the findings suggest that this degree of clinical and longitudinal complexity remained manageable for CHWs, while the study cannot isolate the application’s contribution from training, supervision, medication supply, and other components of the care model.

The strongly positive assessments were accompanied by tensions between standardization and flexibility. Operationally, some CHWs wanted greater control over task timing when participants were unavailable, suggesting that structured follow-up schedules need sufficient flexibility to accommodate circumstances outside the CHW’s control. Clinically, however, standardization remained important, particularly because CHWs were undertaking tasks such as treatment initiation and adjustment. Participants occasionally described recommendations that were difficult to reconcile with care provided elsewhere or with patients’ expectations, particularly when protocol-defined blood-pressure thresholds led to reduction or discontinuation of antihypertensive treatment. These accounts highlight the importance of clear communication across providers, accessible supervision and escalation pathways, and training for CHWs in explaining guideline-based treatment decisions to patients. One participant also reflected on becoming reliant on the application’s confirmation when making decisions; although a single account, it underlines the importance of maintaining CHWs’ clinical knowledge and supervisory support alongside digital guidance, so that decision support augments rather than replaces clinical judgment.

A further theme was accountability. Some participants described the application as lending credibility to their work with patients and colleagues and as ensuring scheduled tasks were genuinely carried out rather than only recorded as complete, suggesting digital decision support may strengthen not only the consistency of care but also its perceived legitimacy and supervision. This visibility was not universally welcomed: a minority saw supervisory oversight as a drawback (6%) or raised data-privacy concerns (5%), and one participant noted that tasks left pending because a participant could not be reached might make it appear the CHW had not worked. Digital traceability can thus strengthen supervision, but performance monitoring must distinguish non-completion attributable to the CHW from tasks unresolved for reasons beyond their control.

Several practical challenges are relevant for wider implementation. The most frequently reported challenge of lengthy questionnaires was attributed qualitatively to research-specific consent and baseline data collection rather than the clinical workflows, so the burden seen here likely overstates that of a platform used primarily for routine care. Charging and intermittent connectivity remained relevant despite offline-first functionality. Although SMS reminders to participants were among the most requested additions, CHWs themselves raised confidentiality and safety concerns about messaging participants on shared phones, illustrating the value of involving frontline users before adding features that may carry social risks. More broadly, these experiences underscore the value of involving frontline CHWs in the ongoing development and adaptation of routine decision-support tools for non-communicable diseases in Lesotho, so that usability and contextual challenges are addressed before wider rollout.

Information continuity across care providers emerged as an additional challenge. Participants described treatment changes that were not always known to CHWs and medication obtained from multiple sources, which could complicate medication reconciliation and create a risk of duplication. These challenges are not specific to the application and would not be fully resolved by digital integration alone, as information on care received elsewhere may remain incomplete and clinical history-taking also relies on patient-reported information. Nevertheless, extending digital workflows across community and facility levels could improve access to shared longitudinal information and reduce avoidable gaps in communication and parallel documentation.

The study has several strengths. All 103 CHWs completed the survey, giving complete coverage and minimizing non-response bias, and the mixed-methods design allowed survey findings to be contextualized and, at times, challenged by qualitative accounts. Notably, the qualitative component elicited criticisms and tensions as well as positive assessments, indicating participants were willing to voice concerns and offering some reassurance against uniformly positive reporting. The evaluation also followed sustained use across 103 villages and three cluster-randomized trials, reflecting repeated field use of a mature application rather than first impressions from a small pilot.

Several limitations apply. The study was conducted within a single research implementation in rural Lesotho, which may limit transferability. Participating CHWs were relatively young and probably more educated than the current national workforce. However, villages were selected independently of CHW characteristics, and replacement CHWs were identified according to Ministry of Health procedures rather than preferentially selected for digital skills. Participants themselves questioned whether older or less-educated CHWs would find the application as usable, and usability may differ in settings with less training, supervision, or infrastructure. Social-desirability bias may have inflated favorable responses, as participants knew the study team had developed the application, although IDIs and FGDs were conducted by a researcher not involved in delivery and the data included criticism. Some survey items used predefined categories that may not capture all experiences; the study assessed perceptions rather than objective performance or diagnostic accuracy and cannot isolate the application’s independent contribution to the trial outcomes. Finally, we did not evaluate the scalability or long-term sustainability of the application itself; the arrangements that routine implementation at scale would require are discussed above.

## Conclusion

The ComBaCaL application was highly usable and valued by CHWs delivering longitudinal hypertension and diabetes care in rural Lesotho, with usability extending from general application functions to complex clinical workflows spanning diagnosis, treatment, medication management and repeated follow-up. This supports the suitability of the open-source CHT Core Framework for complex longitudinal chronic-care applications in this setting and suggests that, with appropriate workflow design, training, supervision and infrastructure, digital decision support can help CHWs deliver guideline-based chronic care. With the clinical evidence from the ComBaCaL trials, these findings support digitally enabled community-based chronic care, including independent first-line medication management, while identifying operational flexibility and integration across health-system levels as priorities for wider implementation.

## Data Availability

The minimal dataset and analysis code are available on Zenodo via https://doi.org/10.5281/zenodo.23730649.

## Acknowledgments

We would like to acknowledge the SolidarMed team in Lesotho and the participating CHWs for their essential contributions to the ComBaCaL project. We are grateful to the team at Medic (www.medic.org) for making the Community Health Toolkit openly available and for the technical support provided. Additionally, we extend our acknowledgements to the students from the Department of Informatics at the University of Zurich, under the supervision of Dario Staehelin and Gerhard Schwabe, for their contributions to developing the pilot version of the application.

## Supporting information

**S1 Fig. Hypertension screening and diagnosis algorithm for people not reporting use of antihypertensive medication.** Blood pressure measurements were taken after selecting the appropriate cuff size, with participants seated after at least five minutes of rest. At the initial visit, blood pressure was measured in both arms, and subsequent measurements were taken from the arm with the higher systolic pressure. The blood pressure value was calculated as the mean of the last two of three consecutive measurements obtained at one-minute intervals. People reporting current use of antihypertensive medication were considered to have hypertension independent of the measured blood pressure value.

**S2 Fig. Diabetes screening and diagnosis algorithm for people not reporting use of antidiabetic medication.** FBG: fasting blood glucose; RBG: random blood glucose. Capillary blood was used for all measurements. Measurements were considered fasting blood glucose if participants reported no caloric intake in the eight hours before the measurement; all other measurements were considered random blood glucose. Type 2 diabetes was defined as (1) self-reported use of antidiabetic medication; (2) an FBG ≥ 7 mmol/l or an RBG ≥ 11.1 mmol/l after a previously elevated FBG or RBG ≥ 5.6 mmol/l; or (3) an FBG ≥ 7 mmol/l or an RBG ≥ 11.1 mmol/l in the presence of all three cardinal symptoms of uncontrolled diabetes (polyuria, polydipsia, and weight loss) in the absence of a clinical history suggestive of type 1 diabetes.

**S3 Fig. Hypertension treatment algorithm applied by community health workers in the intervention villages.** HCTz: hydrochlorothiazide; SBP: systolic blood pressure; DBP: diastolic blood pressure; CVD: cardiovascular disease. ¹De-escalation only if SBP < 120 mmHg and DBP < 90 mmHg. At every visit, the community health worker (CHW) documented current medication, adherence, and blood pressure values. Measurements were taken after selecting the appropriate cuff size, with participants seated after at least five minutes of rest. At the initial visit, blood pressure was measured in both arms, and subsequent measurements were taken from the arm with the higher systolic pressure. The blood pressure value was calculated as the mean of the last two of three consecutive measurements obtained at one-minute intervals. If no contraindications were present, CHWs offered a fixed-dose combination pill containing amlodipine and hydrochlorothiazide, with titration according to the algorithm of the Lesotho Standard Treatment Guidelines. Participants with a blood pressure ≥ 140/90 mmHg under high-dose amlodipine/hydrochlorothiazide, those with contraindications or intolerance to amlodipine/hydrochlorothiazide, and those refusing the medications offered by the CHW were referred to the closest health facility. Treatment was discontinued or de-intensified if systolic blood pressure was < 120 mmHg and diastolic blood pressure was < 90 mmHg. Follow-up visits were scheduled after one month if the dose was adjusted during the index visit, or after two months if the dose was not adjusted. Participants with high CVD risk and no contraindications were offered atorvastatin 10 mg daily. High CVD risk among people with hypertension was defined as a history of stroke or myocardial infarction; diabetes or chronic kidney disease; or being ≥ 50 years old and currently smoking or having a BMI ≥ 30 kg/m². Participants with a history of myocardial infarction or stroke were offered acetylsalicylic acid (aspirin) 100 mg daily.

**S4 Fig. Diabetes treatment algorithm applied by community health workers in the intervention villages.** FBG: fasting blood glucose; RBG: random blood glucose; CDSS: clinical decision support system. Participants not already taking metformin, not meeting referral criteria, and without contraindications were offered metformin initiation at 500 mg/day, with weekly titration to a maximum dose of 2 g/day. In the case of an FBG or RBG < 4.4 mmol/l, the metformin dose was reduced by 500 mg/day. Participants with an FBG > 8.0 mmol/l or an RBG > 9.9 mmol/l who reported good adherence to the maximum metformin dose were referred to the closest health facility for further management. For participants with complicated diabetes, those fulfilling referral criteria, or those with contraindications to metformin, the CDSS prompted CHWs to consult a healthcare professional to guide treatment initiation. Complicated diabetes was defined as use of more than one oral antidiabetic treatment, use of insulin, or meeting referral criteria. Referral criteria were defined as an RBG > 16.7 mmol/l, an FBG > 14.0 mmol/l, or the presence of all three cardinal diabetes symptoms. Contraindications to metformin were defined as severe kidney or liver disease, a previous allergic reaction or intolerance to metformin, or being severely ill.

**S1 Table. Qualitative analysis matrix.** Four individual in-depth interviews and two focus group discussions with 22 community health workers in Butha-Buthe (BB) and Mokhotlong (MKG). The initial analytical framework was informed by domains covered in the quantitative survey and interview guides, with additional codes retained when they arose from the qualitative data. Codes and categories were iteratively refined and organized into four overarching themes. Illustrative quotations are from the English-translated transcripts; ellipses indicate omitted text. IDI, individual in-depth interview; FGD, focus group discussion.

**S2 Table. Item-level responses to the System Usability Scale.** Distribution of responses to each of the ten System Usability Scale (SUS) items among all 103 community health workers. Odd-numbered items are positively worded and even-numbered items are negatively worded; favorable responses correspond to agreement with positively worded items and disagreement with negatively worded items. For each item, the proportion of favorable responses is shown alongside the full distribution across the five-point Likert scale (strongly disagree to strongly agree).

**S1 Appendix. Survey instrument.** Full quantitative survey administered to all 103 community health workers, presented in both English and Sesotho. The instrument covers demographic characteristics, educational attainment, English proficiency, and prior digital experience; the System Usability Scale; items on perceived usefulness, satisfaction, and perceived value adapted from the mHealth App Usability Questionnaire (MAUQ) and the Primary Care Information Project Provider Survey; task-specific ease-of-use items adapted from the Single Ease Question approach; and study-specific items on perceived benefits, challenges, and recommendations for future implementation.

**S2 Appendix. Semi-structured interview and focus group discussion guide.** Guide used for individual interviews and focus group discussions. It explored community health workers’ experiences using the application, perceived benefits and challenges, training and support needs, usability, and the role of the application in supporting the delivery of hypertension and diabetes care.

**S1 Checklist. COREQ checklist.** Completed 32-item Consolidated Criteria for Reporting Qualitative Research (COREQ) checklist for the qualitative component of the study, indicating the location of each item within the manuscript.

**S2 Checklist. GRAMMS checklist.** Completed Good Reporting of A Mixed Methods Study (GRAMMS) checklist, indicating the location of each item within the manuscript.

**S3 Checklist. Inclusivity in global research.** Completed PLOS questionnaire on inclusivity in global research, describing the ethical, cultural, and scientific considerations of research conducted in Lesotho, including local approvals, involvement of local collaborators, community engagement, and plans to share the findings with stakeholders.

